# In vitro characterisation and clinical evaluation of the diagnostic accuracy of a new antigen test for SARS-CoV-2 detection

**DOI:** 10.1101/2021.10.28.21265544

**Authors:** J.J. Montoya, J.M. Rubio, Y. Ouahid, A. Lopez, A. Madejon, A.I. Gil-Garcia, R.J. Hannam, H.R.E. Butler, P. Castan

## Abstract

**Background and aims:** Quick, user-friendly and sensitive diagnostic tools are the key to controlling the spread of the SARS-CoV-2 pandemic in the new epidemiologic landscape. The aim of this work is to characterise a new Covid-19 antigen test that uses an innovative chromatographic Affimer®-based technology designed for the qualitative detection of SARS-CoV-2 antigen. As rapid technology to detect Covid-19, the test was extensively characterised in vitro. Once the analytical parameters of performance were set, the test system was challenged in a test field study. The aim of this study was to evaluate its diagnostic accuracy, as compared by the gold standard RT-PCR and other existing lateral flow tests.

## Introduction

In late December, 2019, a number of patients with viral pneumonia were found to be epidemiologically associated with the Huanan seafood market in Wuhan, in the Hubei province of China. A novel, human-infecting coronavirus, provisionally named 2019 Novel Coronavirus (2019-nCoV) and since renamed SARS-CoV-2, was identified by genomic sequencing (Lu et al., 2020). Spreading primarily through contact with an infected person through respiratory droplets, the incubation period for this virus was confirmed to extend from 2-11 days, causing a series of clinical symptoms that range from mild evidences of runny nose, sore throat, cough, and fever to more severe cases of pneumonia and/or breathing difficulties. In this pandemic context, there has been over 170 M cases of COVID-19 reported worldwide and 3,5 M deaths by the end of July 2021^1.2.3^.

Having to endure a strong lack of molecular diagnostic equipment and testing supplies during the pandemic, molecular biologists have been developing alternatives to help medical professionals accurately diagnose cases of COVID-19, pivoting from RTqPCR from oropharyngeal and/or nasal swabs, to more immediate direct detection of viral antigens in order to generate a combination that had previously proven useful to combat outbreaks of Zika and Ebola^4,5,6^. In this scenario, propelled by the global pandemic, researchers in the UK, have developed a novel biotinylated anti SARS-CoV-2 S1 Affimer® technology that binds to the SARS-CoV-2-S1 protein in anterior nasal swab samples, generating a complex that migrates along a lateral flow strip by capillary action implementing an innovative ultra-sensitive Lateral Flow Device (LFD) based on the interaction of immobilized poly-streptavidin with the migrating complex, via the available biotin label on the anti SARS-CoV-2 S1 Affimer®^7,8^.

This novel anti SARS-CoV-2 S1 Affimer® technology capitalizes on the scientific confirmation that the SARS-CoV-2 2-S1 protein binds to the angiotensin-converting enzyme 2 (ACE2) receptor in humans (Lu et al., 2020) mediating fusion of the viral and cellular membranes (Li, 2016). The LFD tested in this work detects the SARS-CoV-2-S1 protein thorough binding to the novel Affimer® technology, thus targeting a key element in terms of function and structural accessibility, for the S protein assembles to form the trimer spikes protruding from the viral envelope that bring the viral and cellular membranes close for fusion^9,10^.

### Objectives

The starting point was to select the lead anti SARS-CoV-2 S1 Affimer® candidate for efficient binding to the SARS-CoV-2-S1 protein in anterior nasal swab samples and further migration of the complex, based on the characterization of the binding kinetics of the Affimer® reagents towards SARS-CoV-2 S1 protein in a label-free surface plasmon resonance imaging (SPRi) system^11, 12^.

To complete the in vitro characterisation of the selected biotinylated anti SARS-CoV-2 S1 Affimer® within the system, a complete set of tests was performed using purified viral protein. Cross-reactivity and interference tests were completed to generate a specificity profile of the LFD against a) recombinant S1 coronavirus targets and b) common nasal sprays. The analytic Limit of Detection (LoD) was defined, utilizing a) the entire test system from sample preparation to detection and b) spiking inactivated virus (e.g., irradiated virus) into real clinical matrix (e.g., nasopharyngeal (NP) swabs)^13, 14, 15^.

To complete the estimation of the clinical performance (and roughly compare to the in vitro data) a total of 250 samples from volunteers enrolled in test-field epidemiologic studies was analysed, and the sensitivity and specificity were calculated and compared to the RT-PCR data from nasopharyngeal swabs. The studies were completed under the frame of Project SENSORNAS RTC-20176501 in collaboration with MiRNAX Biosens Ltd. and Hospital Carlos III, including a total of 150 negative samples and 100 positive samples, each test documented internally and deposited in agreement to the ISO 15189 norm.

## Materials and Methods

The selection of the lead biotinylated anti SARS-CoV-2 S1 Affimer® candidate(s) was performed attending to the binding kinetics towards SARS-CoV-2 S1 protein. To this end, a set of Affimer® candidates containing a single cysteine were diluted to 5 µM in 10 mM sodium phosphate, 2.7 mM potassium chloride, 137 mM sodium chloride, pH 7.4 (Sigma Aldrich, P4417) and were printed in triplicate to Epoxyde-functionalised SPRi-Biochips (HORIBA, CEp) using an SPRi-Arrayer (HORIBA). The resulting Biochips were incubated (18 °C, 55% RH for 16 h), quenched with 25 mM Ethanolamine (Sigma Aldrich, E9508), 10 mM sodium phosphate, 2.7 mM potassium chloride, 137 mM sodium chloride, pH 7.4 (Sigma Aldrich, P4417), and finally rinsed 4x using 18.2 MΩ.cm ultrapure water (Milli-Q®, Merk-millipore). After assigning spot definitions to the printed Affimer® spots using EZSuite (Ver 1.4.1.68, HORIBA), the biochip was blocked with 1 x Casein Blocking Buffer (Sigma Aldrich, B6429) in PBS-T, 200 µL injection, 25 µL/min for 10 minutes. Reponses for each printed Affimer® spot were then normalised using an injection of 3 mg/mL Sucrose (Fisher Scientific, S/8600/53). Analyte injections of SARS-CoV-2 S1 protein (ACRO Biosystems, S1N-C52H3), were performed at concentrations ranging from 100 nM to 3 nM. Response data for all SARS-CoV-2 S1 analyte injection concentrations was exported from EZSuite and analysed using Scrubber2 (Ver2.0g, HORIBA). Further referencing was performed against the negative reference Affimer® spots, and the association rate constant (ka), dissociation rate constant (kd) and equilibrium dissociation constant (KD) were then determined by fitting using a 1:1 Langmuir model.

In vitro characterisation of the novel anti SARS-CoV-2 S1 Affimer® technology: cross-reactivity and interference testing.

A specificity profile of the system was completed against recombinant S1 coronavirus targets (Table 3) with cross-reactivity observed for SARS-CoV only. In this system, the biotinylated anti SARS-CoV-2 S1 Affimer® reagent binds to the SARS-CoV-2-S1 protein and the antigen-Affimer® complex is then specifically bound to conjugated microparticles. The antigen-Affimer®-microparticle complex migrates along the lateral flow strip by capillary action until it reaches the test line. When contact is made, the immobilized poly-streptavidin on the test line binds the complex via the available biotin label on the Affimer® reagent, whilst the unbound microparticles continue along the lateral flow strip until they reach the control line. Interference testing was also performed with 3 common nasal sprays (Table 4).

In vitro characterisation of the novel anti SARS-CoV-2 S1 Affimer® technology: determination of the limit of detection (LoD).

A pool of (SARS-CoV-2 negative) anterior nasal swab samples inoculated with the inactivated virus (England_02_ strain lot 19/60) was diluted to 10^4^, 10^3^, 5×10^2^, 10^2^ and 5×101 pfu/mL (Figure 1). Confirmatory testing was performed at 1581, 500 and 158.1 pfu/mL giving a pass rate of 20/20 for 1581 pfu/mL and 17/20 for 500 pfu/mL. The final LoD was defined as the lowest concentration resulting in positive detection of 18 out of 20 replicates (90% of all true positives). Assessment of the LFD test was visual. Digital read outs of the line intensity (both test and control) were also generated using a Cube reader.

**Figure 1.**
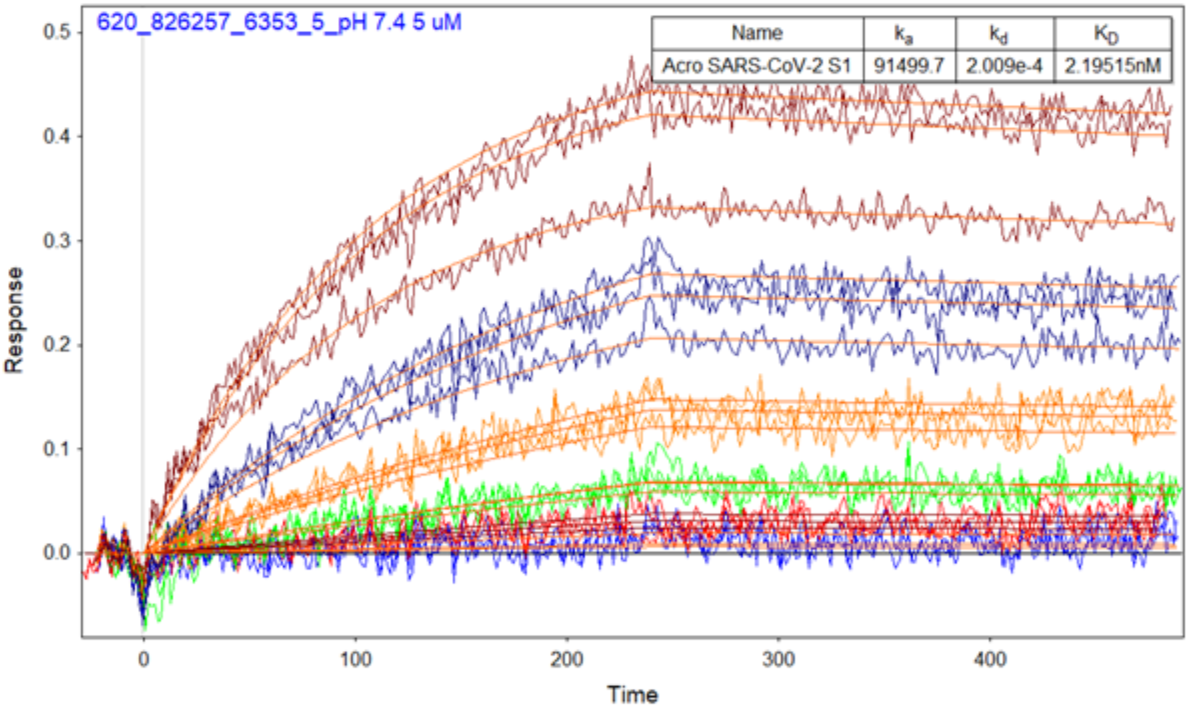
SPRi sensorgrams for triplicate spots of Affimer^®^ candidate 620_826257 with injections of 100 nM – 3 nM SARS-CoV-2 S1 protein, overlayed with 1:1 Langmuir fits estimating Association rate constant (ka) (M-1 s-1), Dissociation rate constant (kd) (s-1), and Equilibrium dissociation constant (KD) (nM).

The estimation of the clinical performance with clinical samples from volunteers enrolled in test-field epidemiologic studies was designed so as to assess the clinical usefulness of the AffiDX® SARS-CoV-2 Antigen Rapid test based on the novel SARS-CoV-2 S1 Affimer® technology, using a nasal swab to confirm the presence of SARS-CoV-2. The studies were completed under the frame of Project SENSORNAS RTC-20176501 in collaboration with MiRNAX Biosens Ltd. and Hospital Carlos III, including a total of 250 samples (150 negatives and 100 positives), each test was documented internally and deposited in agreement to the ISO 13485 norm. comparing the LFD results with results that had been previously obtained from the RTqPCR which was set as the gold standard for detection of SARS-CoV-2. The clinical performance was then estimated within this retrospective structure test-field study collating the positive specimens from consenting patients of any age, gender, or race/ethnicity who presented at the test site with a former PCR confirmation for COVID-19 no older than 4 days. Negative specimens were obtained from consenting patients of any age, gender, or race/ethnicity who presented at the test site with a former negative PCR for COVID-19 no older than 4 days. All LFD tests were performed by minimally trained operators with little laboratory experience who received no previous training on use of the AffiDX® SARS-CoV-2 Antigen Rapid test performed the study test evaluations and were, therefore, representative of the intended users. Study samples: 100 positive (with Ct values ≤30) and 150 negative donors all recently confirmed by RTqPCR were asked to provide the anterior nasal swab which was used to assess the AffiDX® antigen LFD.

**Table 0.** Summary of consumables used for estimation of the clinical performance from volunteers enrolled in test-field epidemiologic studies.

|  |  |
| --- | --- |
| Type of swabs | Beckton Dickinson BBL™ CultureSwab™ dry swab |
| Type of extraction | Extraction protocol - Maxwell® RSC Buccal Swab RNA Kit |
| Type of RTqPCR assay | Applied Biosystems TaqPath COVID-19 CE-IVD RT-PCR |

## Results

Affimer® candidate 620_826257 generated the binding kinetics towards SARS-CoV-2 S1 protein as shown in figure 1, where the SPRi sensorgrams show results in triplicate across the series of injections (100 nM to 3 nM SARS-CoV-2 S1 protein).

Furthermore, Affimer® candidate 620_826257 also displayed good specificity as shown in figure 2, where its positive response to injection of 50 nM SARS-CoV-2 S1 is compared to the negative response to injections of equally prepared other related human coronavirus S1 proteins (MERS-CoV S1, HCoV-HKU1 S1, SARS-CoV S1 and HCoV-229E S1).

**Figure 2.**
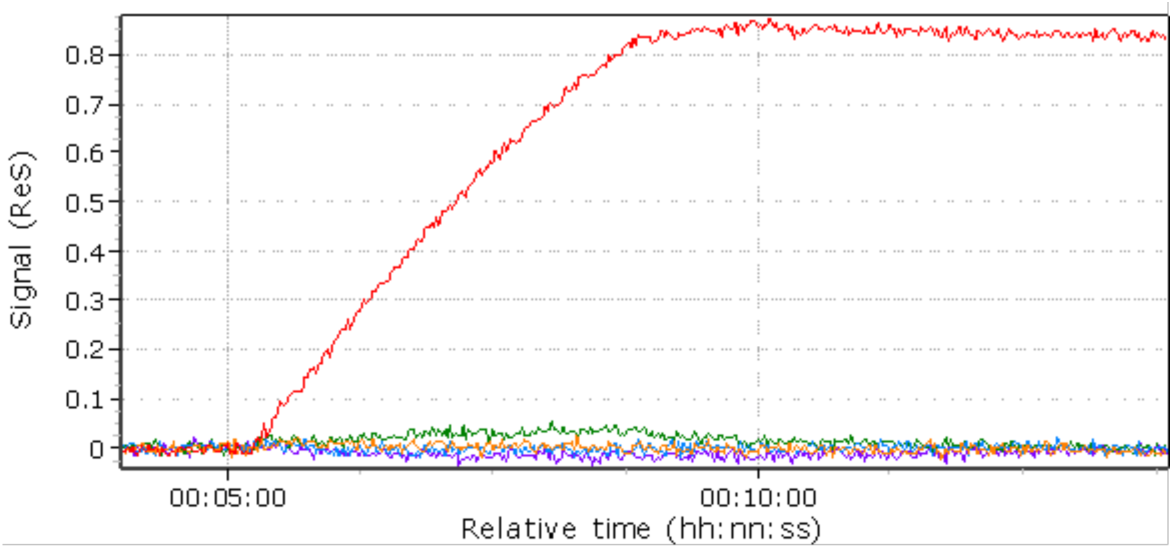
SPRi sensorgrams for Affimer^®^ candidate 620_826257 with injections of 50 nM SARS-CoV-2 S1 and other related human coronavirus S1 proteins: MERS-CoV S1, HCoV-HKU1 S1, SARS-CoV S1, HCoV-229E S1.

**Figure 3.**
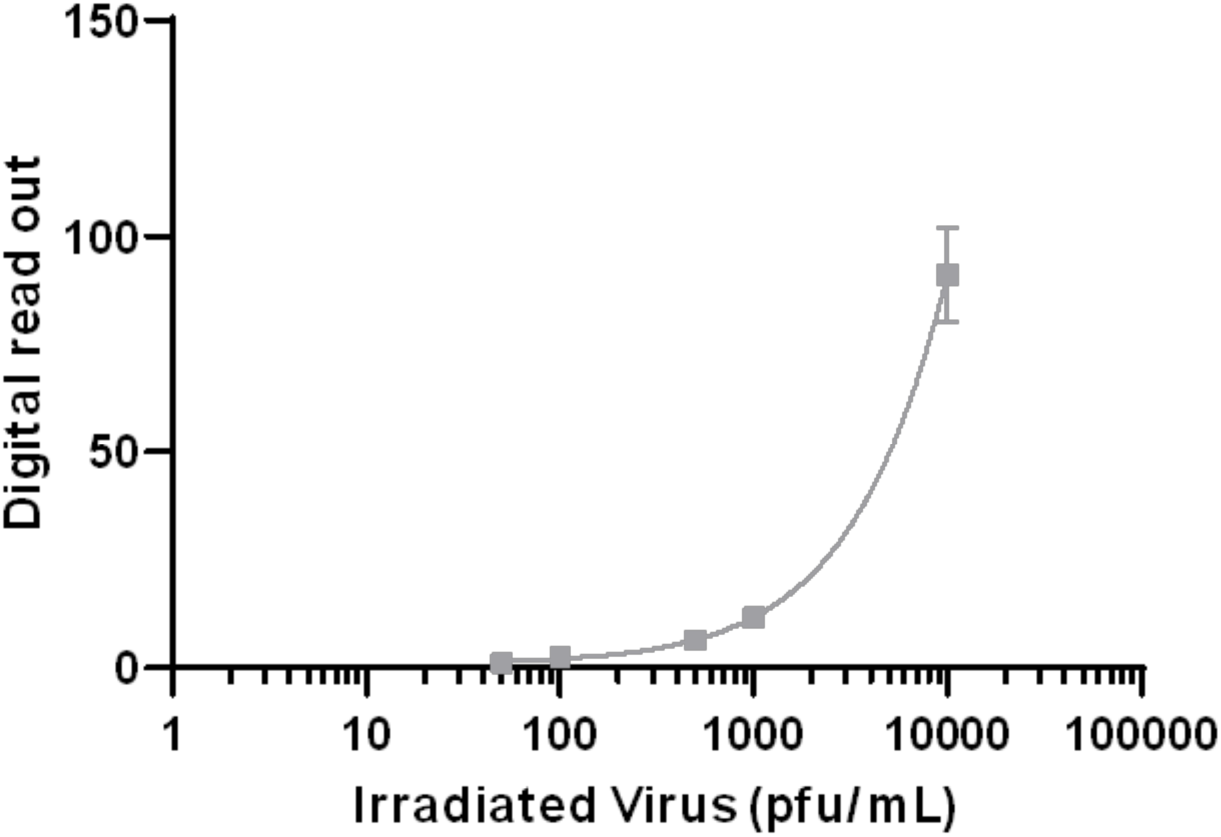
The graph shows digital read outs of the test line intensity against viral titre in pfu/mL across a dilution series prepared using pooled (SARS-CoV-2 negative) anterior nasal swab samples, within which the inactivated virus (England_02_ strain lot 19/60) was inoculated to 10^4^, 10^3^, 5×10^2^, 10^2^ and 5×10^1^ pfu/mL.

In order to define the cross-reactivity and interference testing of the aforementioned LFD system (Materials and Methods) with the chosen Affimer® candidate 620_826257, a specificity profile was completed against recombinant S1 coronavirus targets (Table 1) with cross-reactivity observed for SARS-CoV only. Interference testing was also performed with 3 common nasal sprays (Table 2). Nicorette and Vicks were shown to have no effect on performance of the LFD. Slight interference was observed for Pirinase with reduced intensity of the control and test line. However, no false negatives or positives were observed.

**Table 1.** LFD specificity profile. Cross-reactivity observed for SARS-CoV only.

| **Recombinant S1**<br>**Coronavirus tested at 1µg/mL**<br>**in LFD running buffer** | **Supplier** | **Catalogue**<br>**number** | **Lot number** | **LFD result n=5**<br>**(Positive/Negative)** |
| --- | --- | --- | --- | --- |
| SARS-CoV S1 | Sino Biological | 40150-V08B1 | LC14AP2102 | Positive |
| MERS-CoV S1 | Sino Biological | 40069-V08H | LC14AP2601 | Negative |
| HCoV-NL63 | Sino Biological | 40600-V08H | LC14AP2005 | Negative |
| HCoV-229E | Sino biological | 40601-V08H | LC14MA0701 | Negative |
| HCoV-OC43 | Sino biological | 40607-V08B | LC14MA2003 | Negative |
| HKU1 | Sino Biological | 40602-V08H | LC14AP2908 | Negative |
| SARS-CoV-2 | ACRO Biosystems | S1N-C52H4 | 3532b-204NF1-S1 | Positive |

**Table 2.**
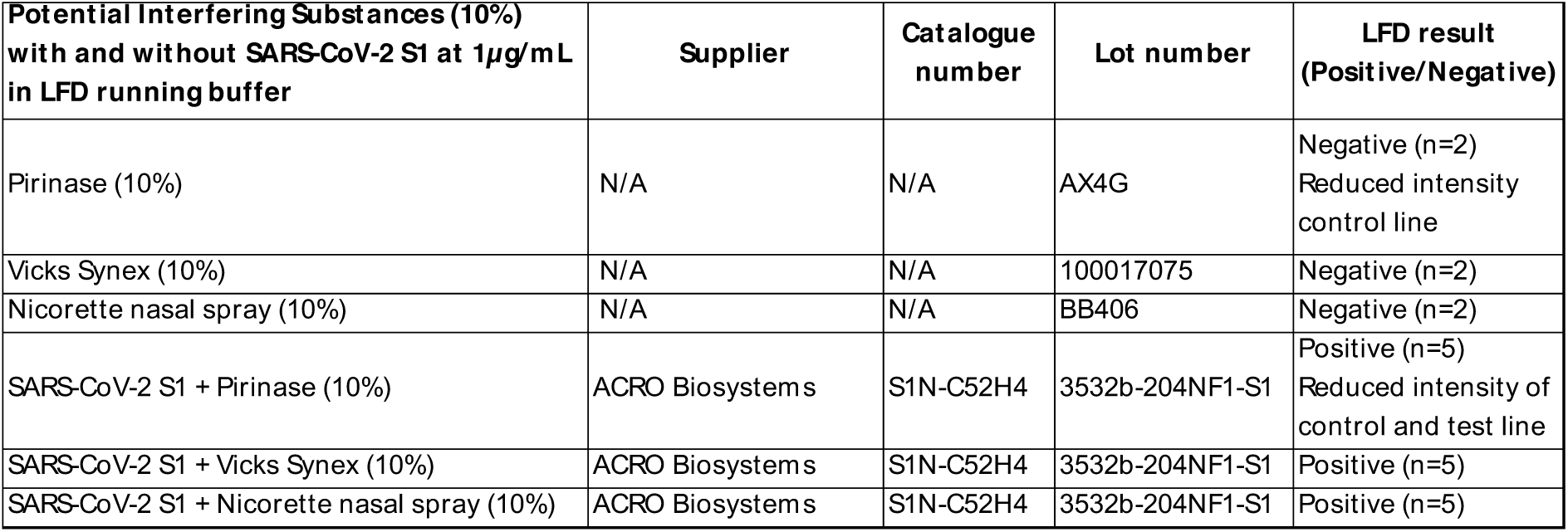
LFD interference testing. Cross-reactivity observed for SARS-CoV only.

The final analytical LoD (defined in the former section as the lowest concentration resulting in positive detection of 18 out of 20 replicates -90% of all true positive-) was confirmed across a dilution series (10^4^, 10^3^, 5×10^2^, 10^2^ and 5×10^1^ pfu/mL). Further testing at 600 pfu/mL was performed providing positive detection for 19 out of 20 replicates (95%), setting this final LoD value.

The clinical performance of the AffiDX® SARS-CoV-2 Antigen Rapid test based on the novel SARS-CoV-2 S1 Affimer® technology was evaluated in the study conducted at different investigational sites in Madrid, Spain as it has been detailed in Materials and Methods. The consenting patients of any age, gender, or race/ethnicity who presented at the test site with a former PCR result for COVID-19 no older than 4 days were tested. RTqPCR data were the standard for comparison of the results from the nasal swab on the LFD with the AffiDX® SARS-CoV-2 Antigen Rapid test based on the novel SARS-CoV-2 S1 Affimer® technology.

150 nasal samples were identified as negative with the AffiDX® SARS-CoV-2 Antigen LFD matching their former 150 negative RTqPCR results. Therefore, there was 100% correlation for the detection of negative samples as shown in table 3, with no false positives observed.

**Table 3.** Correlation between RTqPCR and the SARS-CoV-2 Antigen LFD (Avacta® AffiDX® test) for the negative cohort.

| Determination | Number of Negative identifications | Specificity |
| --- | --- | --- |
| PCR | 150 |  |
| AffiDX® Antigen - Nasal | 150 | 100% |

**Table 4.1 and 4.2.** Correlation between RTqPCR and the SARS-CoV-2 Antigen LFD (Avacta AffiDX^®^ test) for the positive cohort.

| Determination | Number of Positive Identifications | Sensitivity |
| --- | --- | --- |
| PCR | 100 (Ct$\leq 30$) | |
| AffiDx® Antigen – Nasal | 98 | 98% |
| PCR | 100 (Ct$\leq 28$) | |
| AffiDx® Antigen – Nasal | 100 | 100% |

98 nasal samples were identified as positive with the AffiDX® SARS-CoV-2 Antigen LFD from the cohort of 100 positive RTqPCR results with a Ct ≤30, therefore there was 98% correlation for the detection of positive samples with Ct values of ≤30. The 98 samples matched within that cohort had the Ct values of ≤28, therefore there was 100% correlation for the detection of positive samples with Ct values of ≤28. The outstanding specimen had the Ct value ≤30, therefore there was 98% correlation for the detection of positive samples within the cohort.

In summary, the clinical results obtained with the AffiDX® SARS-CoV-2 Antigen Lateral Flow Test based on the novel SARS-CoV-2 S1 Affimer® technology showed a sensitivity of 98% and a specificity of 100% within the cohort tested for a Ct threshold ≤30. Correlation with the reference positive PCR data for Ct values of ≤ 28 showed 100%’s of overlap within the cohort tested. Although the exhaustive comparison of performance against other LFD systems is out of the scope of this work, a sub-cohort of 50 specimens with the Ct values of ≤30, was tested in parallel using other existing SARS-CoV-2 antigen lateral flow tests. There was 98% correlation for the detection of positive samples within that cohort for both Ct values of ≤30 and ≤28, which indicates that the novel Affimer® technology has an edge for detection at lower viral loads (i.e. higher Ct values).

A parallel study under review for publishing will further delve into comparative analysis versus a complete panel of LFDs.

## Discussion

This study is the first to report the journey behind the new AffiDX® SARS-CoV-2 Antigen Lateral Flow Test in terms of in vitro characterisation and clinical performance within a representative cohort. The new AffiDX® SARS-CoV-2 Antigen Lateral Flow Test has shown a strong correlation with the RT-PCR correctly identifying all negative samples and showing no false positives 150/150 (100%). The system’s sensitivity for Ct ≤30 with 98/100 detected (98%) is equally interesting as Ct values ≤28 are considered to be responsible for the vast majority of infectious contacts under an epidemiologic point of view and the system reaches a 100% sensitivity for Ct ≤28 with 98/98 detected (100%)^16^.

Apart from the strong performance data, this new COVID-19 antigen test is an important addition to available tests because the results can be read in minutes, right off the testing card so that the patient gets the information in almost real-time. Due to its simpler design and real usability in terms of clinical performance within the cohort tested, this new antigen test could mark an important advancement in the global fight against the pandemic.

## Data Availability

The studies were completed under the frame of Project SENSORNAS RTC-20176501 in collaboration with MiRNAX Biosens Ltd. and Hospital Carlos III, each test was documented internally and deposited in agreement to the ISO 13485 and all data are available through MiRNAX Biosens S.L.

